# Herd immunity vs suppressed equilibrium in COVID-19 pandemic: different goals require different models for tracking

**DOI:** 10.1101/2020.03.28.20046177

**Authors:** Norden E. Huang, Fangli Qiao, Wang Qian, Ka-Kit Tung

## Abstract

New COVID-19 epicenters have sprung up in Europe and US as the epidemic in China wanes. Many mechanistic models’ past predictions for China were widely off the mark (1, 2), and still vary widely for the new epicenters, due to uncertain disease characteristics. The epidemic ended in Wuhan, and later in South Korea, with less than 1% of their population infected, much less than that required to achieve “herd immunity”. Now as most countries pursue the goal of “suppressed equilibrium”, the traditional concept of “herd immunity” in epidemiology needs to be re-examined. Traditional model predictions of large potential impacts serve their purpose in prompting policy decisions on contact suppression and lockdown to combat the spread, and are useful for evaluating various scenarios. After imposition of these measures it is important to turn to statistical models that incorporate real-time information that reflects ongoing policy implementation and degrees of compliance to more realistically track and project the epidemic’s course. Here we apply such a tool, supported by theory and validated by past data as accurate, to US and Europe. Most countries started with a Reproduction Number of 4 and declined to around 1 at a rate highly dependent on contact-reduction measures.

## 1. Introduction

Almost one hundred years ago, a classic paper published in Proceedings of Royal Society by Kermack and McKendrick (3), entitled “A contribution of mathematical theory of epidemic”, started the tradition of mathematical modelling on the spread of infectious disease among a susceptible population. Current thinking in epidemiology is still deeply rooted in concepts introduced in that paper, some of which are still relevant, while others need to be modified. The mechanistic model they introduced is called the SIR model, for Susceptible-Infected-Recovered/Removed: A susceptible population is infected by the introduction of a few infected individuals. The infected population eventually recovered, and in the process acquiring immunity to the original infectious disease, or dead (removed). An epidemic ends when susceptibles are exhausted as most of the population is immunized this way, achieving “herd immunity”. There have been many variants to this basic model. One common modification is to add an extra population of E for exposed individuals who are not yet infectious, in the so-called SEIR model. Other--- agent-based— models take advantage of the modern computing power to further subdivide the population into many subgroups and even simulate movements of individuals. But basic concepts are similar to the SIR model. These mechanistic models are of critical importance before the outbreak starts, for the models could be used to explore various scenarios for policy decisions on social distancing and lockdown. Once the outbreak starts, a different kind of models are needed—so far not well developed—which are capable of using real-time data to provide the needed parameters for epidemic management, predict various turning points and peak medical resource needs, and monitor the effect of compliance of the quarantine. In this paper, we introduce such a statistical model, supported by theory and validated by observed data.

There are actually two possible end states of an epidemic: one is through “herd immunity” mentioned above when we discuss the mechanistic models, and the other is an unstable state achieved by suppressing contacts among individuals, called “suppressed equilibrium” here, which is pursued by most countries in this pandemic, though many policy makers may not be aware of the distinction between the two. This second state is (parametrically) unstable because if the social distancing measures are relaxed and the businesses reopened, the disease could initiate a second wave, as most of the population has not acquired immunity. Even if the epidemic ends in one country, there could still be subsequent waves of infection by imports from abroad unless there is strict quarantine of cross-border travelers. There is also a third route: to suppress just enough so that the maximum cases of hospitalizations are kept below the maximum capacity of the medical system in each district, delaying long enough either for the population to eventually achieve herd immunity or for a vaccine to be developed for the disease. More people would be infected and more would die in this approach than the suppressed equilibrium, but less than that in the herd immunity scenario, where the hospitals may become overwhelmed. Which approach to take is the difficult decision confronting policy makers at the beginning of an epidemic.

It was reported (2) that UK first contemplated not suppressing the epidemic through lockdowns, fearing that doing so would only lead to a larger second outbreak because most of the population would not have gained immunity. So the plan was to let the epidemic run its course while protecting the elderly. But when shown a model prediction (4) that such a “do-nothing scenario” would lead to 500,000 deaths and 81% of the population infected, policy makers changed course and imposed strict counter measures. This is an important and proper role for a model, to prompt policy actions to combat the spread of the disease. Once the outbreak started, the accuracy of the mechanistic model predictions cannot be verified as the forecast forever changed the course of the epidemic in UK. Health officials in Sweden didn’t believe in models and decided to pursue “herd immunity” starting 12 March.

The number of people that will have to be infected before achieving herd immunity depends on how contagious the disease is. The 16 March report of Ferguson et al.(4) assumed an infection rate, expressed in terms of Basis Reproductive Number *R*_0_ of *2.4*. For US it predicted that 81% of the population would have to be infected in this do-nothing scenario, or about 250 million, resulting in 2.2 million dead. Later updates in the 30 March report of Flaxman et al. (5) suggested that *R*_0_ should be above 4 for European countries studied. Estimating this number will be one of the tasks in the present work so that one can evaluate what it entails for the herd-immunity approach. We will later show, directly from data, that this estimate of *R*_0_ ~4 also holds for the US, and in fact approximately so for every country we examined. So over 90% of the US population would need to be infected before herd immunity could be achieved. COVID-19 turns out to be much more contagious than originally thought. See also (6).

Since a “suppressed equilibrium” is achieved in a very different manner than that for the “herd immunity”, our estimate of the end date of the epidemic as a consequence of contact suppression is not based on the number of susceptibles, *S*, approaching a small critical value (i.e. when most of the population is infected, hence acquiring immunity), but the daily new cases *N*(*t*) approaching zero and remaining so for two incubation periods. For prediction purpose, the date when the *N*(*t*) is near zero is estimated by 3 standard deviations from its peak. These two quantities, the peak and the standard deviation, can be extracted from the data as the epidemic is developing. Our estimate of the end of the epidemic is earlier, usually significantly so, because it does not depend on a high percentage of the population having been infected to achieve herd immunity.

For South Korea, the epidemic in that country ended with just 0.02% of its population infected. Wuhan, the epidemic ended with less than 0.5% of its population infected, both less than 1% of that required to achieve herd immunity as predicted by most mechanistic models. Even if these numbers are multiplied by a factor of 10 (as some in the media suggested that the Wuhan data may be an underestimate), it is still one order of magnitude less. Furthermore, it is generally accepted that South Korea’s data are good. Indeed, the situation in these countries represents early examples of the “suppressed equilibrium”. Because of its much lower number of deaths, such an end state is a goal that most countries have decided to pursue, despite the enormous toll on the economy due to the much reduced business activity for the two to three months that it would take to achieve it.

The above-quoted numbers for Wuhan and South Korea are for the confirmed cases, and do not include the asymptomatic infected. Recently (in early April) about 3,330 individuals in Santa Clara County in California were tested for antibodies to the COVID-19 virus in their blood (7). 50 were tested positive (meaning that they were exposed in the past to the virus), yielding a crude prevalence rate of 1.5%. When weighed by demographics and extrapolated statistically to the whole county’s population, it was calculated that 2.8% of the population could have been infected, with a 95% confidence range of 1.3-4.7%. These numbers, less than 5%, are also much less than what is required to achieve herd immunity, in a county where the epidemic is waning at the time.

In pursuing the “suppressed equilibrium” and monitoring the progress towards it, a second type of statistical models is required. This type of models should reflect the contact-reduction measures already in place and the degree of compliance by the population in each region. Unlike China and South Korea, many European countries imposed these measures in stages, and so the contact rate among the population was reduced in a time-dependent way. In US, even before the epidemic is over, some states are starting to reopen businesses. Only a real-time, data driven model can reflect these changes. Such a model should be based on sound epidemiological principles. Prediction based on a purely statistical model without an epidemiological foundation, even though using real-time and past data, is akin to watching the daily stock market fluctuations and performing “technical analysis” to ask when the peak is for an investor to time a sell order. A prediction could be made but the uncertainty would be so large that it is just likely for the prediction to be right as for it to be wrong.

Each type of models has its strengths and weaknesses. For the mechanistic model, such as SIR and SEIR or agent-based versions, a key parameter, the infection rate, is not known for an emerging disease such as SARS-CoV-2, and this has been a source of difficulty with predictions using such models. For the second type of models, especially the purely statistical models without epidemiological basis, it is not known which quantity of the epidemic is predictable. For example, there have been many empirical models based on the assumption that the progression of daily cases follow a Gaussian “epidemic curve” in time, starting with the early model of William Farr in 1840: “Law of Epidemics” in his second annual report to the Registrar General of England and Wales (8). Lacking the epidemiological mechanism that Kermack and McKendrick (3) later proposed, the “law” simply reflected Farr’s conviction that the observed deceleration of the rate of increase of infected would not lead to an impending catastrophe but to a crest and then accelerated decline. The latest is that of the Institute of Health Metrics and Evaluation (IHME)(9). It turns out that fitting three parameters that define a Gaussian to a short time series and then using that Gaussian to predict the peaks of the epidemic is an ill-posed problem(10). The uncertainty for next-day prediction is near 100%, and should increase further for predictions a few days out (11). In this work we pay special attention to which epidemiological property is predictable, and we quantify the uncertainty of our prediction.

The search for the correct “geometry of epidemic curves” has a long history in statistical modelling. Farr’s law is purely descriptive. Farr did not realize that his epidemic curve is Gaussian but nevertheless his descriptive law of second ratios could be used for prediction, though not very accurate. For example (see (12)), if *x*_1_*,x*_2_*,x*_3_*,x*_4_*,x*_5_*,x*_6_*…*. are the successive weekly incidence (i.e. new cases) or mortality, his law says that the ratio of successive ratios of these numbers is a constant:

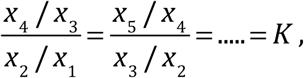

which is less than 1. That is, there is a constant deceleration of the rate of growth of the cases. After measuring this constant from the early weeks’ data, future incidence values can be predicted. It was John Brownlee in 1907 (13) who realized that the above formula, when logarithm is taken—turning the ratio of ratios to difference of differences—is a finite difference form of the second order time-derivative of log *x* being a negative constant (12):

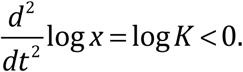

Integrating twice and then taking exponentiation lead to *x*(*t*) being a Gaussian form. Brownlee thought this normal form for the epidemic curve is a fundamental law in epidemiology, but his proposed explanation for the declining growth of the incidence of an epidemic as due to decreasing “infectivity” was not well-received by epidemiologists at the time.

Brownlee (13) provided examples of several epidemics showing that there was fore-aft symmetry in their epidemic curves. For COVID-19, we find that the epidemic curve for Wuhan, China follows a Gaussian, with near fore-aft symmetry, but that for US has a rapid rise but slow decline, definitely not Gaussian. While it may be possible that without human intervention, a solitary outbreak may follow a Gaussian curve, in the modern era of contact suppression, the epidemic curve is shaped by such interventions. We shall explain Wuhan’s shape as due to the fact that the contact suppression measures were consistently imposed throughout the course of the outbreak, while in the case of US, its states and the populace, were relaxing earlier measures on the aft side of the curve, when the new cases declined, creating a fore-aft asymmetry. Therefore, one should take into account that in the modern era, as countries pursue a “suppressed equilibrium” at great economic cost, there is a tendency in countries with decentralized state governments to relax the countermeasures to various degrees once the disease crested, giving rise to subsequent waves of infection. [A second reason is that the US data is an aggregate of data for different epicenters, with staggered recovery. A third reason for the asymmetry may be artificial: In many countries there is usually an increase in the testing of the population as the production of the test kits and facilities to process them ramp up after the initial shortage. Therefore it is to be expected that there would be more cases found later in the course of the epidemic than in the beginning.]

Brownlee rejected the idea of herd immunity, that the epidemic’s decline was due to “an exhaustion of susceptibles, enshrined 20 years later in the SIR model of Kermack and McKendrick (3). His alternative, “infectivity” idea was based on the thinking that the decline was due to “the loss of infecting power on the part of the organism”(13), and that this biological property of the pathogen (“organism”) should follow some fundamental law. This biological property of the virus has not been observed in the current COVID-19 pandemic, and does not appear to be a factor. However, Brownlee’s idea can be resurrected by modifying the definition of “infectivity” to include social factors, since how many people one infected individual can infect, as measured by the Effective Reproduction Number, *R_t_*, depends on the product of the number of persons contacted during the infectious period and the probability of the contacted person contracting the disease. After implementation of contact-reduction measures, we can actually see from the data (in section 2) the decline of this measure of “infectivity”. Furthermore the decline is steeper in countries that have the more stringent contact-reduction policies and implementation. Although both the mechanisms of loss of susceptibles and decrease in “infectivity” are likely at play, with the extremely small percentage of the population infected in the current pandemic, the second mechanism appears to be the dominant one as countries strive to achieve the “suppressed equilibrium”. Given this situation, model predictions of the decline of the epidemic based the number of susceptibles decreasing, as in SIR and SEIR models, may be missing the main cause for the observed progression of the disease in the current pandemic.

Brownlee’s idea, with modification expressed above, can be cast in a mathematical form as:

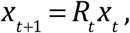

where *x_t_* is the incidence (new cases) at time *t*, and *x_t+_*_1_ is the incidence one infectious period later. An infection period is the duration an infected person remains infectious. *R_t_* is defined earlier as the number of people one infected individual would infect during the period when he is infectious. If *R_t_* is a constant, *R_t_ = R*_0_, the solution to the above finite difference equation is:

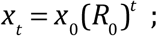

the solution is an unimpeded exponential growth (since (*R*_0_)*^t^ =* exp{*t* log *R*_0_}) for the relevant case of *R*_0_ *>* 1. Brownlee(14) commented that such an epidemic form is contrary to the facts: "The assumption that the infectivity of an organism is constant, leads to epidemic forms which have no accordance with the actual facts.” With *R_t_* as a decreasing function of time, which we find is actually the case in section 2, the above solution becomes Gaussian-like. Specifically, if *R_t_* decreases by a factor *q <* 1 after each period (12, 14), due to a “loss of infecting power”, i.e. *R_t_ = R*_0_*q*^(t−1)^, then the solution is Gaussian:

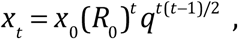

(since 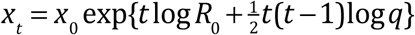, noting log *q <* 0).

The exponential growth from (*R*_0_)*^t^* is eventually overtaken by the more rapid decrease of 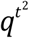. Note that in this argument, no mention is made of the decrease of the number of susceptibles; this is not needed when the decrease is so small compared to the population as a whole.

This paper is organized as follows: We first introduce the relationship between the net infection rate and the Reproduction Numbers. The epidemiological basis for our model is discussed. Then, we will present a suite of prediction tools for epidemic management. We will also provide a summary of the COVID-19 inferred epidemiological characteristics for various countries in Asia, Europe and US. Finally, a discussion and conclusion will be given. The theoretical support is given in the appendix.

## 2. The net infection rate and the Reproduction Numbers

Before we discuss our prediction model, we first discuss diagnosing a key parameter used as input in most mechanistic epidemiological models, the infection rate, or equivalently expressed as the *Basic Reproduction Number* or the time-dependent *Effective Reproduction Number*. Using real-time data, we diagnose it for different countries in the world, which actually reflects the underlying influence factors.

We define in general the *net infection rate α*(*t*) as the time-varying exponential growth rate of the active infected cases (15):

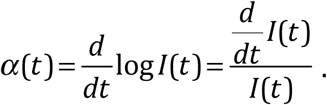

The active infected number, *I*(*t*) is given by the equation that describes its rate of increase as the daily new infected cases *N*(*t*) minus the daily recovered/removed case, *R*(*t*):

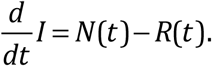

The dead (“removed”) is included in *R*(*t*) in our calculations. The peak number of active infected cases is a key parameter in the planning for hospital resources. This turning point, denoted by *t_p_*, can be located in a local-in-time manner by when *R* starting to exceed *N*, without first accumulate the data in time to find *I*(*t*). Maximum demand for hospital resources occurs at its peak, and not at the peak of *N*(*t*), although the latter is a more commonly reported quantity. *R*(*t*) is not a factor in the initial rise of the outbreak, nor is it needed to explain why the rise slows and eventually crests. However in the waning phase, when the new cases are smaller, the rising recovered cases need to be taken into account to determine *t_p_*.

In traditional mechanistic models, such as the SIR model(3), there is also a time-dependent net infection rate, which at *t=*0, when the population is completely susceptible, is related to the *Basic Reproduction Number R*_0_. See ref (16) for a discussion of the complexities associated with this key parameter. We will not be using the SIR model but it is useful to relate our general definition to what is traditionally used. The equation for *I* in the SIR model is:

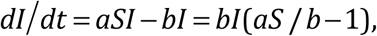

where *aS*(*t*) is the infection rate and *b* is the recovery/removal rate.

Therefore 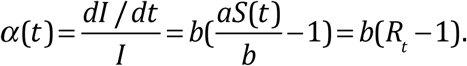.

So for the SIR model 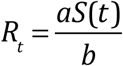. So as the population of the susceptible *S*(*t*) decreases---“exhaustion of the susceptibles” as hosts for the disease—the Effective Reproduction Number decreases. More relevant for our discussion is the possibility that the infection rate *a* could change as a result of contact reduction measures in place.

Initially when the whole population is not yet infected, the Basic Reproduction Number is 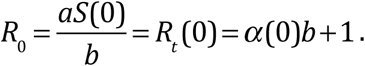.

The above-defined time-dependent net infection rate generalizes this concept to be independent of the SIR or other models: If in the course of an epidemic, *α*(*t*) is positive, the number of infectives will grow exponentially, reaching a peak number of infectives when *α*(*t*) = 0 at *t = t_p_*, which is a critical turning point mentioned above. Then the total number of active infectives will decrease exponentially. In terms of *R_t_ =α*(*t*)/b +1, if this number is greater (less) than 1 the total number of active infectives will grow (decrease) at time *t*. We will here use *α*(*t*) directly. *R_t_* however is the more watched quantity by the mainstream modelers (16). It can be calculated from the net infection rate, but will require a parameter *b*, the recovery rate, which may be different for different regions. Furthermore, many countries do not keep adequate records on those who recovered, and so there is an uncertainty in estimating *b*. In Figure 2, *R_t_* is obtained by estimating this parameter as *b* ≈ 1*/σ_R_ ≈* 1*/σ_N_*, where *σ_R_* is the standard deviation for the distribution of the daily recovered and *σ_N_* is that for the daily newly infected numbers. These parameters are calculated for each country (see Table 1). *R*_0_ is obtained from *R_t_* in the initial period, before there is significant recovered population.

**Table 1.** Summary of the predictions made on May 5, 2020. **Notes**: Countries with “ *”sign after a country name indicates that the epidemic is still developing, although some of the particular event might have happened. The number in the parenthesis is the actual data. A“+” sign indicates the latest value in the particular case that is still developing and that particular event has not happened yet. The ‘#’ indicated the result might be questionable, for the lack of recovered case numbers. IPM= Infection per Million of Population. FPM=Fatality per Million of population.

| | $t_p$<br>(r) | $t_N$<br>(r) | $t_D$<br>(r) | $AIC_{peak-K}$<br>(r) | $TIC_{\infty-K}$<br>(r) | $AFC_{\infty-K}$<br>(r) | $\sigma_N$<br>Days | CFR<br>--% | IPM<br>--Per M | FPM<br>--Per M |
| --- | --- | --- | --- | --- | --- | --- | --- | --- | --- | --- |
| China | 2-21<br>(2-17) | 2-7<br>(2-13) | 2-17<br>(2-14) | 44.4-54.1<br>(58.0) | 87.7-163<br>(84.0) | 3.8-6.6<br>(4.6) | 8.79 | 5.53 | 59.98 | 3.31 |
| China<br>ex-Hubei | 2-11<br>(2-12) | 2-5<br>(2-4) | 2-16<br>(2-13) | 8.99-9.04<br>(8.94) | 15.5-26.1<br>(15.8) | 0.14-0.23<br>(0.13) | 8.49 | 0.79 | 11.82 | 0.09 |
| South<br>Korea | 3-15<br>(3-14) | 3-2<br>(3-1) | 3-19<br>(3-29) | 7.08-8.35<br>(7.49) | 8.84-16.2<br>(10.8) | 0.18-0.30<br>(0.26) | 6.31 | 2.36 | 209.42 | 4.94 |
| Italy* | 4-23<br>(4-21) | 3-30<br>(3-21) | 3-31<br>(3-28) | 114-115<br>(108) | 203-399<br>(213+) | 24.8-42.1<br>(29.3+) | 12.18 | 13.76+ | 3526.7+ | 485.3+ |
| Ger-<br>many* | 4-4<br>(4-7) | 3-31<br>(3-27) | 4-14<br>(4-15) | 50.7-69.5<br>(70.8) | 144-309<br>(167+) | 6.86-14.0<br>(7.0+) | 9.62 | 4.19+ | 2014.6+ | 84.35+ |
| Spain* | 4-22<br>(4-27) | 4-6<br>(3-26) | 4-7<br>(4-1) | 114-129<br>(105) | 274-495<br>(248+) | 28.1-48.4<br>(25.6+) | 10.44 | 10.32+ | 5316.9+ | 548.5+ |
| USA* | 5-12<br>(NA) | 4-19<br>(4-9) | 4-22<br>(4-16) | 1400-1474<br>(943+) | 1520-2410<br>(1204+) | 90-140<br>(71.1+) | 12.21 | 5.9+ | 3649.5+ | 215.3+ |
| UK* | 6-30#<br>(NA) | 4-20<br>(4-10) | 4-20<br>(4-30) | 11789#<br>(165+) | 249-392<br>(195+) | 33-62<br>(29.4+) | 14.21 | 15.1+ | 2932.2+ | 442.5+ |

For an emerging disease such as SARS-CoV-2, there is not enough statistics for estimating the true infection rate (e.g. the parameter *a* in the SIR model). Models usually assume a value of *R*_0_ or perform scenario calculations for a range of values of *R*_0_.

Figure 1 shows the net infection rate for several countries. Since the official data that we use include only the confirmed cases (“cases” for short), and these tend to have more serious symptoms that require hospitalization, the reported *I* cases are commonly referred to as *total hospitalizations*. The peak of total hospitalizations is closely watched by hospital administrators and policy makers. *α*(*t*) is commonly referred to as *rate of hospitalization*. Its inverse gives the e-folding time in days for the cases in an outbreak. A value, of say *α* between 0.3 and 0.4, where most countries cluster in the initial period, implies an e-folding time of about 3 days (doubling time of 2 days). The much higher values *α* for many regions in the beginning of our data record may not be due to indigenous disease infection. See later discussion.

**Figure 1.**
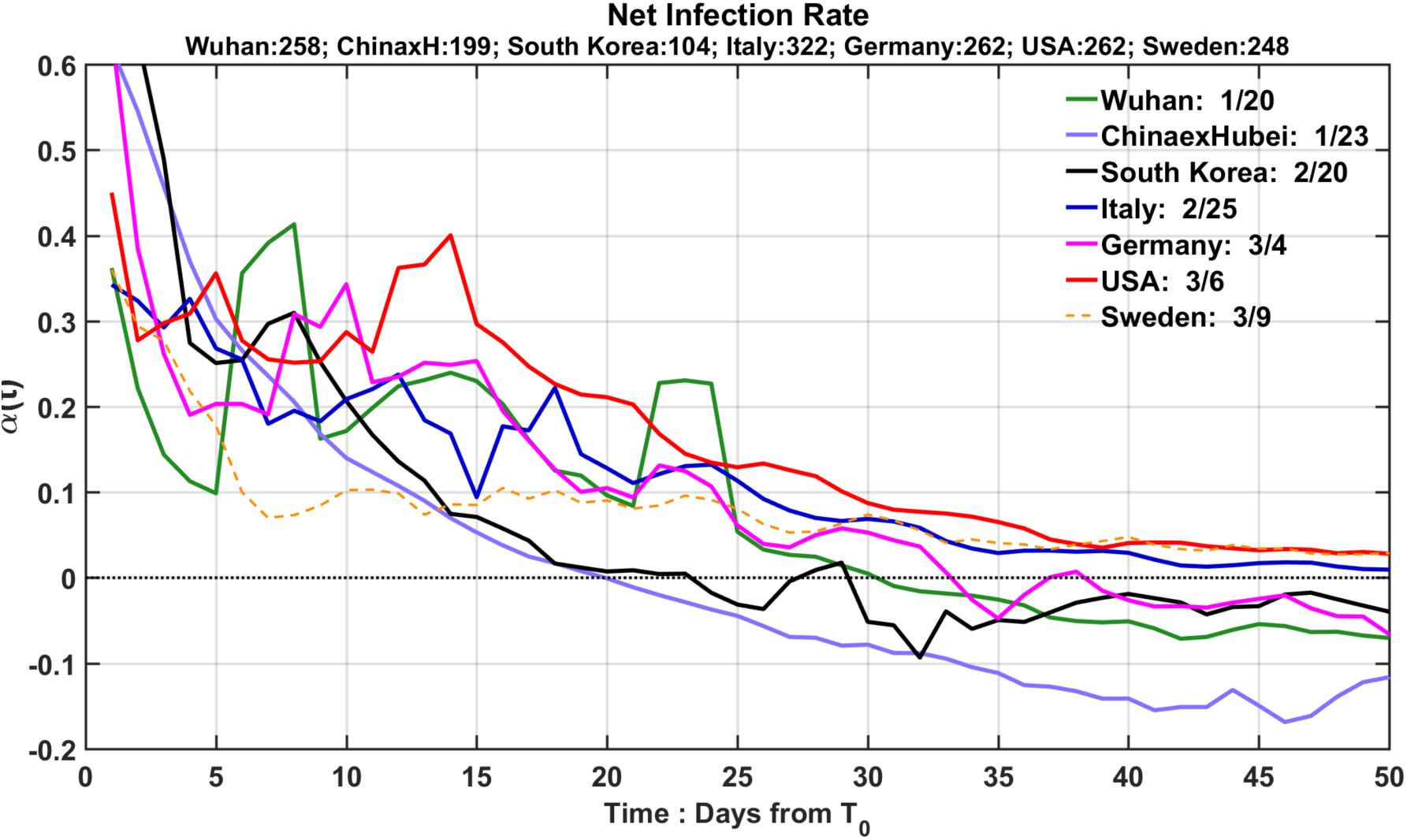
The time-dependent net infection rate (in units of 1/day) as a function of time starting on the date (listed in the inset) when the newly confirmed case number first exceeds 100 for each region. To obtain the actual calendar date, add the dates on the horizontal axis to the starting date indicated in the inserted legend. The number of confirmed cases on the starting date is listed at the top. Three day averaging on the raw data has been used.

**Figure 2.**
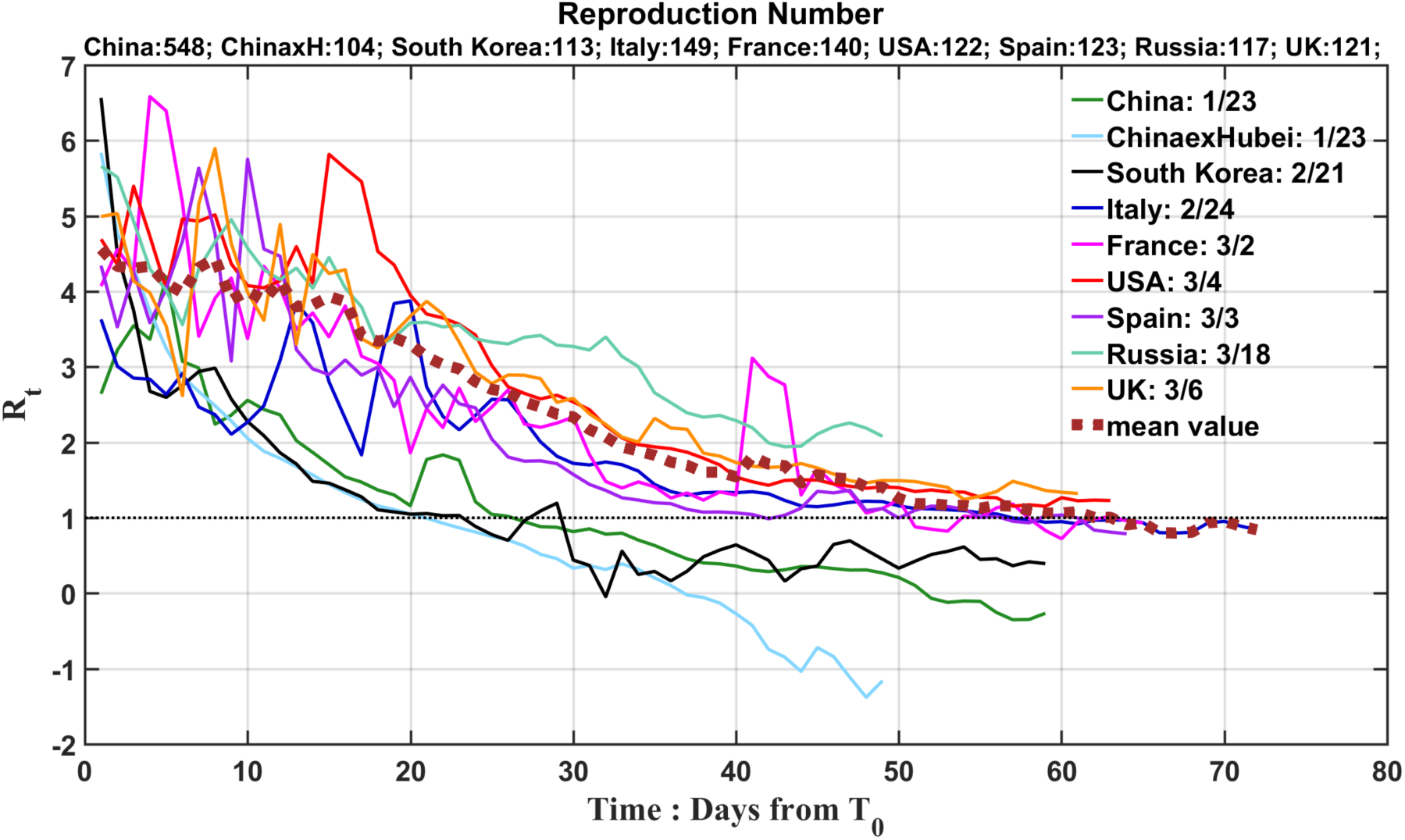
Effective Reproduction Number for each country or region. The horizontal axis denotes days since day 0 (the corresponding calendar date is given in the inset), which is the starting date for our calculation. This date is determined by the threshold that the accumulated number of infectives first exceeds 100. The actual number for each region on that day is listed at the top. The thick dashed curve is the average of the curves for USA and European countries, including Russia.

The time for different countries is aligned in Figure 1 to begin the time series when each region reached 100 new cases. This way, the progression of the epidemic in each country can be compared. Figure 1 reveals the effects of different policy measures each country adopted. First, South Korea and China exHubei have similar net infection rates (until past their respective turning point); both are much lower than other countries. In the case of South Korea, the government identified early that its epicenter of the epidemic was at church gatherings in the city of Daegu and North Gyeongsang province, where 90% of the initial cases were found. Specifically, a confirmed COVID-19 patient was reported to have attended the Shincheonji Church of Jesus services twice on February 9th and 16th. Then aggressive contact tracing was pursued. After the turning point, South Korea soon experienced some second wave episodes, which were successfully contained. These two regions’ rigorously implemented contact reduction and aggressive pursuit of ‘test-trace-treat’ measures led to them being the extreme examples of the “suppressed equilibrium”.

Germany and Italy have similar exponential growth rates of the net infected case numbers, both slightly higher than Wuhan. More surprisingly, US has the highest exponential net infection rate, 1.5 times that of Germany and Italy and twice that of Wuhan. This can be attributed to the fact that US so far does not have a nation-wide lockdown, and Europe has had partial lockdowns in phases. Germany took a week longer than Wuhan to reach its turning point, while US will take weeks longer than Germany. China outside Hubei reached its turning point early, in fact 9 days earlier than the epicenter, Wuhan. This fact is significant, for it is qualitatively different than many mechanistic model predictions, which had the epicenter achieving its turning point 1-2 weeks earlier than China outside Hubei (17), probably based on the herd immunity concept.

The net infection rates for China outside Hubei and South Korea are more monotonic than other regions shown. This is due to the fact that there was not a piece-meal imposition of social-distancing measures, unlike other western countries. The strict measures were imposed and enforced throughout the course. [For Wuhan, the large spike on day 23(12 February) was due to a change in the diagnostic criteria from a positive nucleic acid test to chest scans, booking in one day more than 13,000 cases.]

The case of Sweden needs a special explanation. The epidemic in Sweden initially grows with an e-folding time of around 3 days, in line with other countries. Then on 12 March the government announced that because of limited resources, it no longer would test for the COVID-19 infection, except for those with serious symptoms already in the hospitals who furthermore are also in the high-risk group. As a result the new cases took a nose-dive on that day, leading to an artificially low net infection rate of 0.1, implying a 10 day e-folding time. The denominator in the calculation for *α*(*t*) is *I*(*t*), which is an accumulated quantity, and includes those who tested positive prior to 12 March under more liberal criteria, and so this situation leads to a flat, low level of *α*(*t*) just above 0. It would eventually cross 0 with a large enough death numbers. Sweden’s policy decision to pursue “herd immunity” (while protecting the elderly) has been touted as a viable and perhaps preferable approach to those of other countries in their pursuit of “suppressed equilibrium”. It only encouraged those over 70 to stay home and banned visits to the nursing home and gatherings with over 50 people, while business, stores, restaurants and kindergarten through grade nine are open. The success or failure of this approach cannot be evaluated by the incomplete data. Intriguingly, based on the recorded death number, it shows that Sweden’s toll is 5 and 11 times that of its neighbors Denmark and Norway, respectively. The population in each of these neighbors is half that of Sweden.

Figure 2 converts Figure 1 to show *R_t_* for each country. It shows that *R_t_* clusters around 4 for all countries in the initial period. Because of the problems for the data in the initial period, the curves cannot be extended back in time to deduce *R*_0_. But based on *R_t_* a few days later, *R*_0_ for COVID-19 should be around 4, similar to that for SARS. It was originally thought that COVID-19’s *R*_0_ was between 2.0 to 2.5 (18), seemingly much less contagious than SARS at 4 (19). It is also much morecontagious than the 2009 swine flu pandemic, caused by the H1N1 virus, whose *R*_0_ was estimated(20) to be 1.4 to 1.6.

In deducing the Reproduction Numbers we should not count the large spike for China outside Hubei on day 4. That increase was not due to indigenous transmission, for most of the initial cases were imported from Hubei. This should not be used to infer the Reproduction Number. Similarly for South Korea in the first few days shown.

As is in Figure 1, the decrease of *R_t_* from 4 to 1 for different countries reflects different level of contact-reduction measures adopted and enforced. With China outside Hubei and South Korea sloping more steeply than Europe than US. The behavior of these numbers for the European countries are rather similar to the model results of Flaxman et al.(5) of Imperial College (their Figure 2), who imposed these measures in their model on the dates they were actually imposed. With their emphasis on more accurately predicting the mortality, when *R*_0_*~*4 is used in their SEIR model the modeled death numbers are close to the reported deaths, but the number of infected in the model is an order of magnitude higher (Figure 2 in (5)). This is reasonable and consistent with our earlier statement that the data of infected cases do not include the asymptomatics or those with mild symptoms who were not tested, but reflect those with more serious symptoms that required hospitalization, and therefore more prone to die.

UK’s record for the recovered is almost nonexistent, and what is available shows that the recovered number is only a few percent of the deaths, which does not appear to be reasonable. Without the recovered in the data, UK’s *R_t_* hovers above 1. There appears to be a similar situation in some other countries, such as Italy. Therefore, the behavior of *R_t_* in the later stages of the epidemic (in the neighborhood of the turning point) is probably not correctly depicted by the data shown for these countries. Nevertheless, in the initial period, when the number of recovered is small, the data shown can be used to estimate *R_t_* and *R*_0_.

SEIR model was also used to deduce the Reproduction Number by Institute of Disease Modeling (21) in an effort to monitor the effect of social distancing measures adopted near Seattle (King County, Washington). They found that *R_t_* was reduced from 2.7 to 1.4. Since it was not below 1, the Institute’s report advised continuing the measures in place. Since the report rate *p*, that is, the ratio of the number of reported cases vs. the true infected number, was unknown, the authors assumed a range of values and obtained, *R*_0_ ~2.7 ± 0.9. One can see from this application of the Reproduction Numbers how important it is to monitor in realtime the progress of policy measures to determine whether it is time to relax the measures in place. And also how difficult it is to infer these numbers.

The report rate *p* is the ratio of the confirmed cases to the true infected numbers. If it were known, the case numbers can be divided by *p* to yield the true infected numbers. Because it is largely unknown at the present time, it creates the aforementioned uncertainty in estimating Reproduction Numbers using models. However, since our net infection rate *α*(*t*) is a ratio of the derivative of *I* and *I*, dividing each by *p* does not change the ratio. We therefore recommend using the net infection rate for real-time monitoring instead of the Reproduction Number. The assumption is that the report rate has not changed in time, at least not during the two-week period prior to *t* that the accumulation takes place for the denominator (see Appendix). This same consideration is also partly behind our later using the ratio of new cases to recovered cases for prediction.

The problem with the data for Sweden is that in addition to the change in the report rate, the record for the recovered is not available.

This discussion also highlights the difficulty of using mechanistic models for realtime monitoring. In addition to not knowing the report rate *p* to compare model output with the reported case numbers, a key parameter needed in the mechanistic models, the infection rate *a*, is largely unknown for the emerging disease. There is a large population of asymptomatic, untested and unreported infectives. This infected population is nevertheless infectious and produces some of the infected cases reported. It would usually lead to an overestimate of the infection rate *a* when only the reported cases are used in the estimation. Our tools do not need to use an infection rate for prediction.

## 3. Epidemiological basis

After establishing the relationship between the net infection rate and the Reproduction Numbers, we briefly summarize the main ingredients to establish the epidemiological basis of our model. Details of our model are given in the Appendix. The model is used here only to infer general properties of an outbreak, and to discover which properties can be predicted.

Based on epidemiological theory, the infected population is governed by the Von Foerster equation in an age-structure population model (see (22)), where “age”, *s*, is days since first infected. The infection at age 0 is governed by the “birth” dynamics, the infection dynamics, and death due to natural causes is ignored. An infected individual is assumed to be infectious in an age-dependent way until *T*, when the individual is either cured or dead, in either case no longer belonging to the population of the infectives, where *T* is the mean recovery/removal period. The solution to this partial equation (via the method of characteristics) yields certain rather general results.

### Conservation Law

For *t > T*

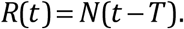

The distribution of the newly recovery/removal follows that of the newly infected with a time delay of *T*. When applied to hospitalizations, it says that those who are admitted to the hospital either recover after a mean hospital stay of *T* days, or dead after a similar number of days. A more complicated relationship holds for *t < T*.

Since hospital stay acts like a filter for *R*(*t*), the profile for *R* is slightly wider than that for N. Appendix takes this into account.

*R*(*t*) here actually consists of two parts: the recovered individuals *R_c_*(*t*) and the removed (dead) individuals, *D*(*t*)*: R*(*t*)*= R_c_*(*t*)*+D*(*t*).

### Validation after the fact

This fundamental relationship can be validated statistically with data, provided the data is long enough. This is one of the ways the mean recovery period *T* is determined statistically from data, but it is not practical in the early phase of the epidemic.

Since *R_c_*(*t*) and *D*(*t*) are a subset of *R*(*t*), the time lag relationship should also hold. Additional insight can be gained by looking at them separately. Figures 3, obtained using the longest data from China, South Korea and Italy during the COVID-19 pandemic, shows that *N*(*t*) and *R*(*t*) are highly correlated: with correlation coefficients all over 0.9 when both distributions are smoothed with 3-point boxcar. The mean time delay of the correlation is denoted by *μ*, which can be interpreted as a statistical mean of *T*. The spread of the cross-correlation is measured by its standard deviation, *σ_c_*, its value is also given in the figure. The lag time (and standard deviation) of *R*(*t*) for China is 19 days (*σ_c_*=17.7 days) for South Korea is 23 days (*σ_c_*=19.8 days) and for Italy is only 10 days (*σ_c_*=21.2days). Due to their low Case Fatality Rate (CFR), there is practically no difference between *R*(*t*) and *R_c_*(*t*) for China and Korea. But for Italy, which has a high CFR, there are differences among the constituting parts. Why Italy’s lag time between *R*(*t*) and *N*(*t*) is shorter than China and South Korea needs a deeper examination, and it does not necessarily mean the shorter the better. The lag time of *D*(*t*) for China is 7 days, for Korea is 17 and for Italy is only 3 days. The short survival time for patients in the hospital is an indication that the Italian hospitals were overwhelmed and allocation of ventilators was selective to those more likely to survive. This mortality component reduces the overall time for recovery/removal for Italy to 10 days.

**Figure 3.**
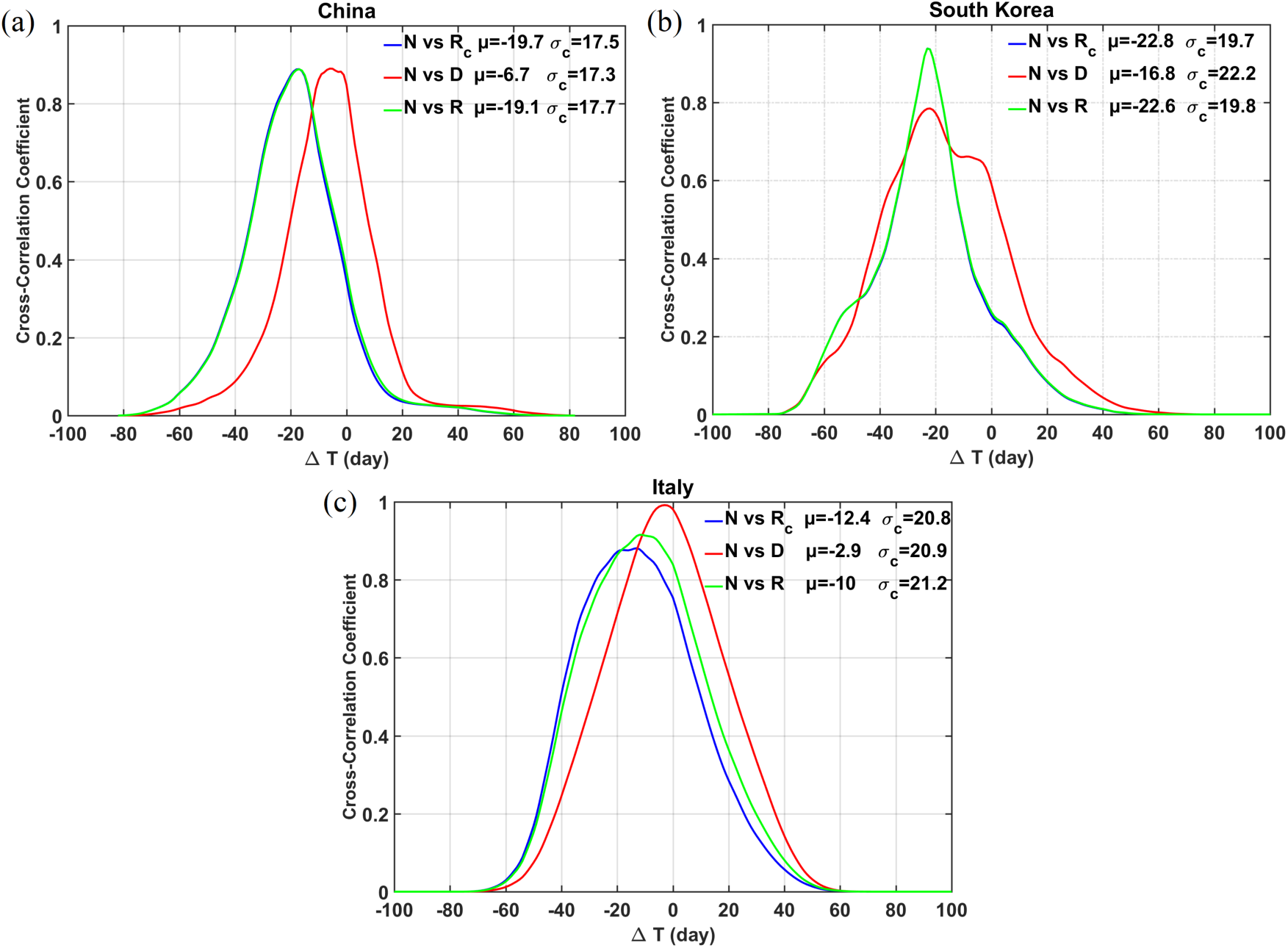
Lagged cross-correlation of *R_c_*(*t*)*, D*(*t*) and *R*(*t*) with *N*(*t*) for China, South Korea and Italy. Collectively or individually, the conservation law and time lag relationship are validated. The separated *R_c_*(*t*) and *D*(*t*) reveal the stress the regional hospitals had experienced.

### Epidemic curve

Furthermore, the solution to the model equations (in Appendix) shows that the time-dependent profile of *N*(*t*), sometimes referred to as “the epidemic curve”, is determined by the “age” distribution, and the method of characteristics in the solution converts that “age” distribution into a time profile. The details of the “age” distribution depend on the “birth” process and on how long it has been since the outbreak first begins. We assume that we do not mix data from different epicenters (the homogeneity assumption). Focusing on a single outbreak, which starts at *t=*0, we examine the “age” distribution at a time *t_B_* much longer than the initial incubation period. Then we assume there exists a full spectrum of age with *s* between 0 and *T* in a Gaussian-like form. To the right of the peak in the “age” distribution, it is easy to understand that those who are “older” should be less in number because they were infected during an earlier stage of the epidemic. As the epidemic grows exponentially, there are more and more “younger” infectives. To the left of the peak in “age’ distribution many infectives are at various stages of incubation. Those who are newly infected may be much large in number, but since they have not yet developed symptoms, they are less likely to be tested, hospitalized, and contribute to the “case” record. For Covid-19, the peak infectiousness occurs in a period just before and after the onset of symptoms, around 5-6 days since first infected(23). The above discussion gives an epidemiological justification for a Gaussian-like distribution of the “age” distribution of the “new cases”, more appropriately called “new hospitalization per day”. It is consistent with the argument given in the Introduction in favor of a Gaussian-like epidemiological curve. It should be pointed out that historically, for obvious reasons, asymptomatics were also not included in the “incidence”, or “mortality”, which were used by Farr to obtain his law of second ratios or by Brownlee to obtain his normal curves.

With the possible exception of South Korea, in most countries there could be multiple seeding of the outbreak occurring at slightly different times. These staggered series of outbreaks merge in a continuum in the data for that country, leading to a standard deviation in the new cases that is wider than that from the “age” distribution. This is a complication we need to take into account.

Also, in many countries, pressure mounts for policy makers to relax the contact-reduction measures when case counts pass the peak and are declining. In some countries where the restrictions are gradually lifted, we should expect a long tail in the epidemic profile, which is therefore not symmetric with respect to the peak. This external influence to the original expected progression of the course should be monitored and adjustment to predictions made in real time. As will be discussed in subsequent sections, our model has the ability to adapt to these changes in its role as a monitoring tool. Nevertheless, we find, after the fact, that predictions for the various turning points are still accurate even without taking into account the changes. It turns out that, consistent with the above discussion, the relaxation of contact-reduction measures, which lengthens the standard deviation of the new cases, is only significant in the later stages of the course, and can be ignored before the peak, but the prediction on the evolution after the peaks on quantities such as the end of the epidemic and the total number of infected is likely not accurate unless the changes are taken into account. (This has not been done in Table 1.)

## 4. A suite of tools for tracking the epidemic

Based on theoretical considerations, we developed a suite of tools for monitoring the evolution of the epidemic. They can be used to predict the timing of several turning points and the number of infectives associated with each. These include *t_N_*, the peak of the daily new cases; *t_R_*, the peak of daily recovered/removed cases, and *t_p_*, the turning point where the active infected cases (AIC), or total hospitalizations, is a maximum. This is the point of maximum strain on the hospital resources. It needs to be closely monitored to keep the AIC below the hospital capacity. We also estimate the date when the epidemic ends and the country can be reopened, *t_c_*.

A more accurate and robust prediction tool is based on the ratio of *N*(*t*) and *R*(*t*), designated as the *NR* ratio. This ratio also alleviates to some extent the problem related to the data of reported cases being a fraction *p* of the true numbers, as *p* cancels out in the ratio. Unfortunately, some countries, such as UK and Sweden, do not keep adequate record of *R*(*t*), and many country do not keep a rigorous standard, which could be detect by the low case recovery rate, indicating the violation, or leakage, of the conservation law. For these countries a less accurate method, in the sense of having larger error bars, can still be used when only information on *N*(*t*) is available. These are described below. We have used the data from countries that have the longest records for COVID-19 to verify properties of these methods.

### 4.1 Log of NR ratio

We define the *NR* ratio as

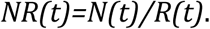

At *t_p_, NR=*1.

We show in Figure 4, using the data of the epidemic for COVID-19 for the longest records available in some countries, that the logarithm of *NR*(*t*) lies on a straight line, with small scatter, passing through the turning point *t_p_*. And data for various stages of the epidemic, from the initial exponential growth stage, to near the peak of *AIC*, and then past the peak, all lie on the same straight line. The intercept with *logNR=o* yields the turning point. This line, obtained by linear-least-square fit, is little affected by the rather large artificial spike in the data on 12 February for China, because of its short duration and the logarithmic value. That reporting problem is necessarily of short duration because, on the date of definition change, previous week’s cases of infectives according to the new criteria were reported in one day. After that, the book is cleared, and *N*(*t*) returned to its normal range.

**Figure 4.**
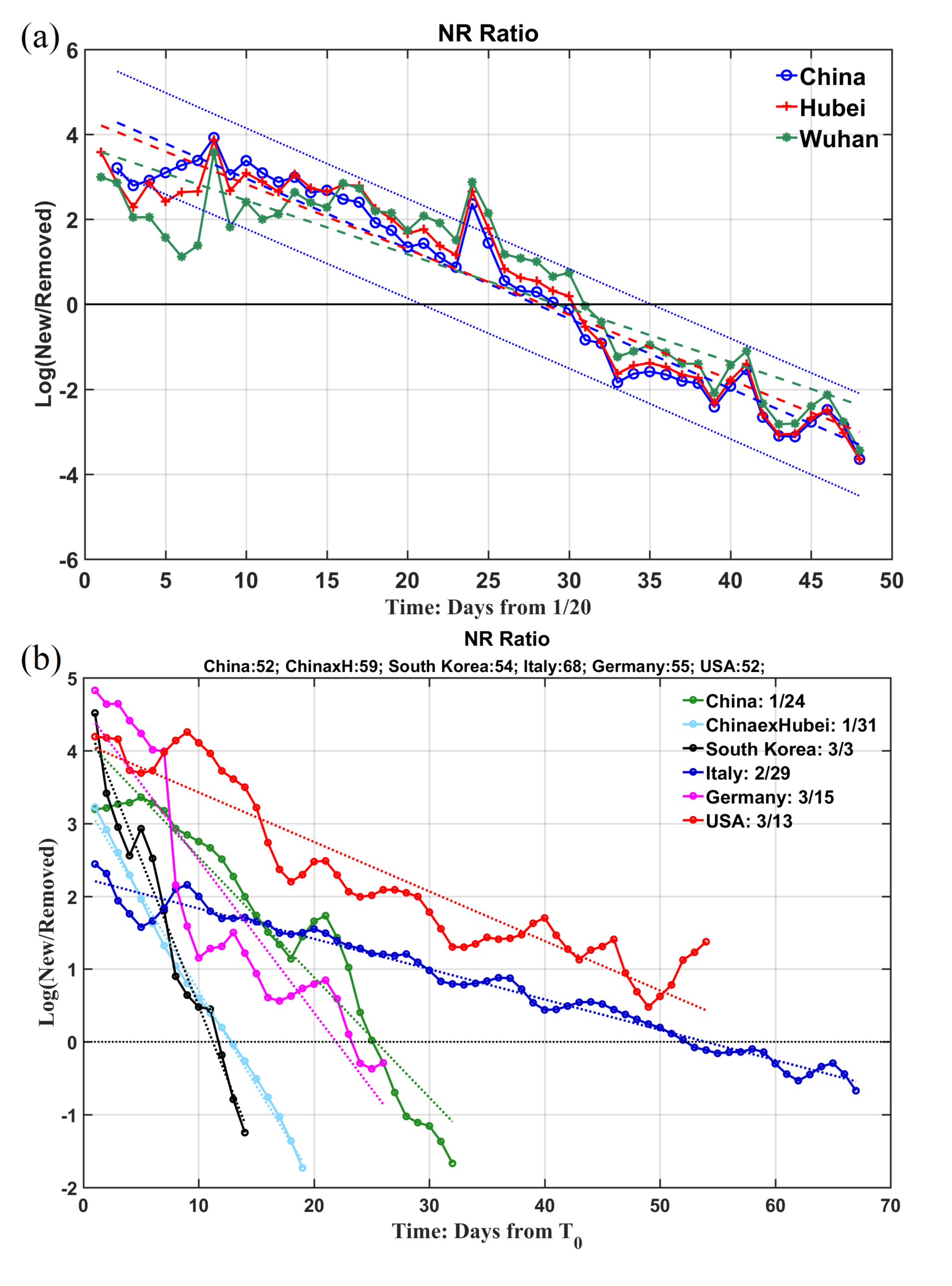
Natural Logarithm of the ratio of daily newly infected to newly recovered/removed. They lie on straight lines with some small scatter. The straight line obtained by linear-least squares fit is in dotted line with the same color for each country. (a) For China, the slopes of the lines are almost the same but with different intercept; the trend lines cross zero (the black solid line) at different time for different regions indicating different peaking time for *AIC*. The epicenter Wuhan (green) has latest turning point than its province Hubei (pink), which has a later turning point than China as a whole (cyan). As a measure of confidence of the linear-least square fit, the 95% confidence limit for China is given in the figure. Similar confidence limits for other regions have been calculated but are not shown for the sake of clarity of the presentation. (b) Comparing different countries. The time is aligned by plotting the *NR* ratio only when the recovery/removal case numbers first exceed 50 (the actual numbers on that day are listed at the top), and *T*_0_ is that calendar date, listed in the inset. The figure in the bottom panel included the China cases again, to facilitate comparison with other countries, except the data used were smoothed by a 3-point boxcar filter in the lower panel.

It would be interesting to understand why the empirically determined log *NR*(*t*) lies on a straight line, and what determines its slope. See Appendix for a theoretical support. It is shown that, if the distribution of the new cases is Gaussian-like, the natural logarithm of the *NR* ratio, should be a linear function of time throughout the course of the epidemic. The slope of the line is 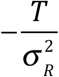.

*σ_R_* is the standard deviation of the recovered case distribution and is close to that of the newly infected case, *σ*_N_, twice of which measures the duration of the epidemic in that region. This expression is still valid even for a sigma that is changing in time as a response to changing social contact measures.

A comparison of the Logarithm of *NR* ratio for several countries is given in Figure 4 (b). A steeper slope is associated with an early turning point, and also a predictor for a shorter duration of the epidemic. The shallowest slopes in Figure 4 (b) was for Italy, where the enormous pressure strained their medical system to the limit, resulting in the largest *σ_R_* value, and one of the highest case fatality rate in the world, at more than 12%. Germany and China have similar slopes. For China outside Hubei, the slope is steepest and the turning point reached 9 days earlier than Wuhan. South Korea’s slope is even steeper due to that countries early action. As a result, Italy took a full month longer to reach its turning point than Germany and China, and more than 40 days longer than South Korea.

### 4.2 Predictability

Since the logarithm of *NR* ratio lies on a straight line passing through the turning point of *AIC*, it would be interesting to explore if the turning point, *t_p_*, can be predicted by extrapolation using data weeks before it happened. Extrapolating a straight line is much more practical than other more involved curve fitting algorithms some other groups have adopted. How far in advance this can be done appears to be limited by the poor quality of the initial data, when *R* is small and highly fluctuating. Figure 5 (a) shows the results of such predictions for China, and 5 (b) for Italy. It is a hind cast since the truth is now known (see Figure 8 for how it is determined). The horizontal axis indicates the last date of the data used in the prediction. The beginning date of the data used is 24 January for all experiments for China. Prior to that day, data quality was poor and the newly recovered number was zero in some days, giving an infinite *NR* ratio. For China outside Hubei, the prediction made on 6 February gives the turning point as 14 February, two days later than the truth. A prediction made on 8 February already converged to the truth of 12 February, and stays near the truth, differing by no more than fractions of a day with more data.

**Figure 5.**
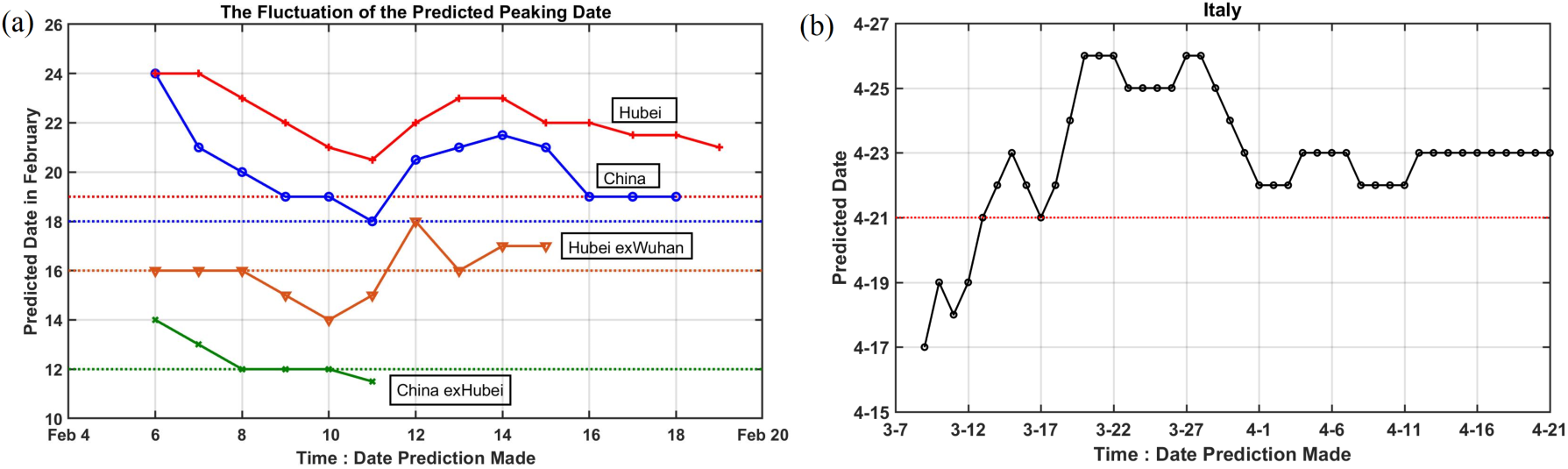
(a) Prediction of the turning point in *AIC* for different regions of China by extrapolating the trends in logarithm of *NR*. The horizontal axis indicates the date the prediction is made using data from 24 January to that date. The vertical axis gives the dates of the predicted turning point. Dashed horizontal lines indicated the true dates for the turning point. (b) Prediction of the turning point in *AIC* for Italy. Data used for all predictions starts on 29 February. The first point shown predicts the turning point to occur on 19 April (4 days early) 6 weeks in advance using 9 days of data from 29 February to 8 March.

The huge data glitch on 12 February in Hubei affected the prediction for Hubei, for China as whole, and for Hubei-exWuhan. These three curves all show a bump up starting on 12 February, as the slope of *N*(*t*) is artificially lifted. Ironically, predictions made earlier than 12 February are actually better. For example, for China as a whole, predictions made on 9 February and 10 February both give 19 February as the turning point, only one day off the truth of 18 February. A prediction made on 11 February actually gives the correct turning point that would occur one week later. At the time these predictions are made, the newly infected cases were rising rapidly, by over 2,000 each day, and later by over 14,000. It would have been incredulous if one were to announce at that time that the epidemic would turn the corner a week later. Even with the huge spike for the regions affected by the Hubei’s changing of diagnosis criteria, because of its short duration the artifact affects the predicted value by no more than 3 days, and the prediction accuracy soon recovers for China as a whole. For Hubei, the prediction never converges to the true value, but the over-prediction is only 2 days.

Uncertainty associated with prediction using this method for China is shown in Figure 4(a). The uncertainty is a few days (the 95% confidence level is ± 5 days) and is usually somewhat larger than the accuracy. A prediction can be found later to be accurate but at the time it was made it may be equally likely for the prediction to be a few days earlier or later. The large uncertainty is again seen to be caused by the 1,4000 bump of new cases in one day on 12 February due to the change in definition For applications to other countries and to future epidemics without a change in the definition of the “infection” to such a large extent, we expect even better prediction accuracy and smaller uncertainty.

This can be seen in the prediction for Italy. The error of predicting the turning point 3 weeks in advance is only 1 or 2 days. In fact a prediction can be made 6 weeks in advance with an accuracy of 5 days or less.

The prediction for US as a whole is less accurate (with errors up to 10 days) because its data is an aggregate of different epicenters. More accurate predictions can be made by treating each state separately. This is not done here because although the data for new cases and deaths are available for each state, recovered data are not individually available. It is also not accurate for UK because its data for recovered may be suspect. See Table 1.

### 4.3. Derivative of Log of *N*(*t*) and *R*(*t*)

Interestingly, the derivative of log *N*(*t*) or log *R*(*t*) also lies on a straight line, as shown in Figure 6 (although the scatter is larger as to be expected for any differentiation of empirical data). Moreover, the straight line extends without appreciable change in slope beyond the peak of *N*(*t*), suggesting that the distribution of the newly infected number is approximately Gaussian-like at least up to that point. For an exponential function, the derivative of its logarithm being a linear function of time is highly suggestive of a general type of distribution including Gaussian-like or Rayleigh-like. The recovery time *T* can be determined as *t_R_-t_N_*, where *t_R_* is the peak of *R*(*t*) and *t_N_* is the peak of *N*(*t*). These two peak times can be obtained by extending the straight line in Figure 6 (a) to intersect the zero line. This predicted result can be verified statistically after the fact by the lagged correlation of *R*(*t*) and *N*(*t*). If the distribution is indeed Gaussian or even approximately so, the slope in Figure 6 should be proportional to the reciprocal of the square of its standard deviation, *σ*, as:

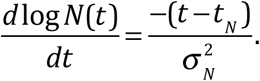

Similarly result holds for the daily number of recovered, *R*(*t*).

**Figure 6.**
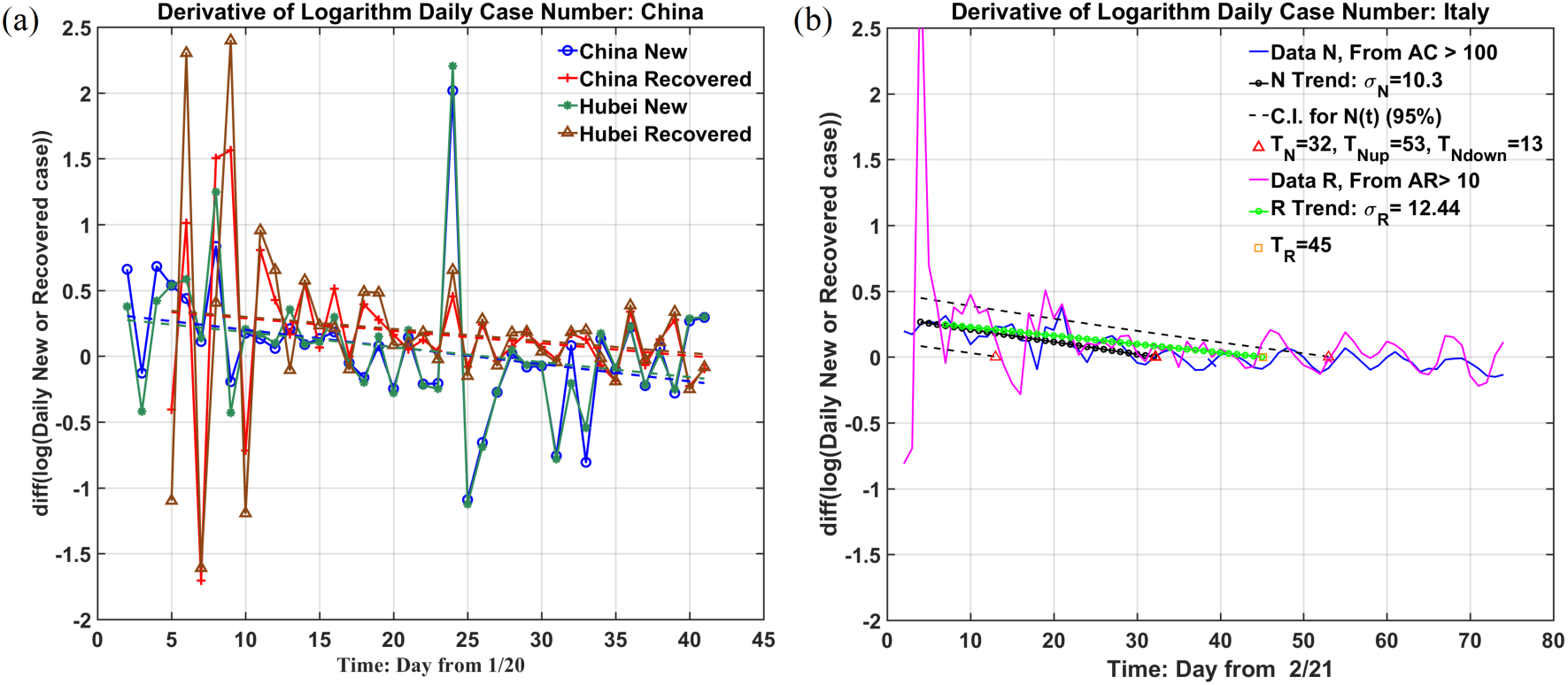
(a). The derivative of the logarithm of daily newly infected or recovered for China and Hubei. Notice the clear separation of the new and recovered cases and also the subtle difference of their slopes. The zero crossings of the trend line give the peak dates of the new and recovered case respectively. And the slopes give an estimate of *σ* values. (b). Same as (a) but for Italy. Best fit lines for *N*(*t*) and *R*(*t*) are shown, and 95% confidence limits are shown for *N*(*t*) only. Italy’s data appear to have a 7-day periodic oscillation, probably caused by a reporting issue. AR indicates accumulated R.

For countries without an adequate record of *R*(*t*), this method can still be used for *N*(*t*). We can obtain *t_N_* and *σ*_N_. *T* and *t_p_* cannot be obtained, but can be estimated roughly as *t_p_ = t_N_ + T* /2 (see Theory), using *T*~20 days applicable to countries with similar medical systems.

### 4.4 The number of deaths

Because of public attention, predicting the number of deaths has been the main emphasis for some models. There is also the belief that death numbers are more reliable than case numbers, although that is not necessarily true. Although death number by itself could not satisfy conservation law, death is a subset of the newly confirmed cases; therefore, the death case distribution should follow the new cases with a time delay. In fact, this delay time could provide a measure of the efficacy of the medical system, as explained earlier.

In Figure 7, we present the cases for Italy, USA and UK.

**Figure 7.**
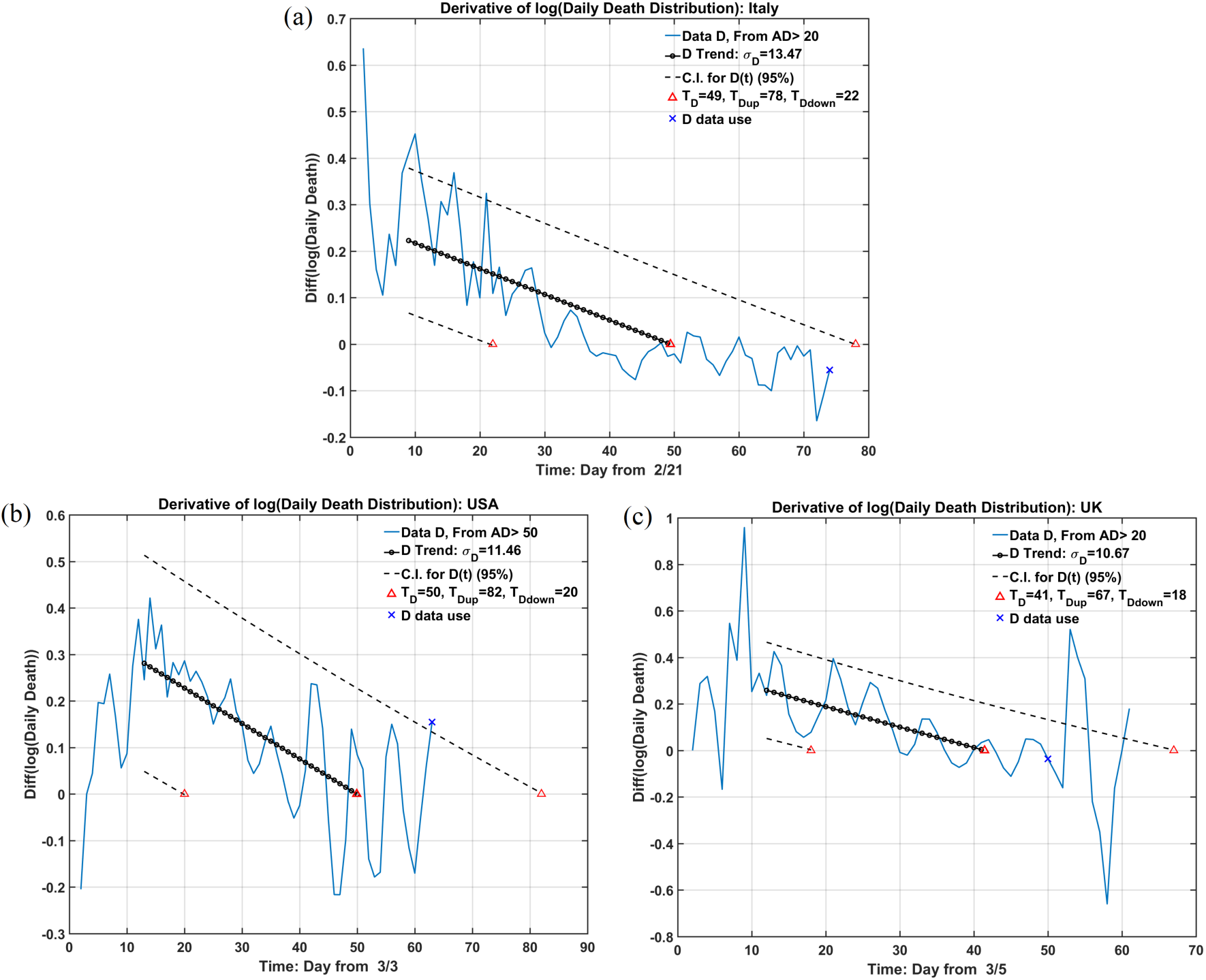
**(a), (b) and (c)**. Logarithm for death distribution for Italy, USA and UK. Similar to 4.2, the derivative of logarithm of *D*(*t*) follows a straight line approximately, indicating that its distribution is also Gaussian-like, similar to *N*(*t*). As a consequence, the peak of *D*(*t*) occurs at the intersection of that straight line with zero and could be predicted in advance, although in the three countries shown this peak has already occurred.

**Figure 8.**
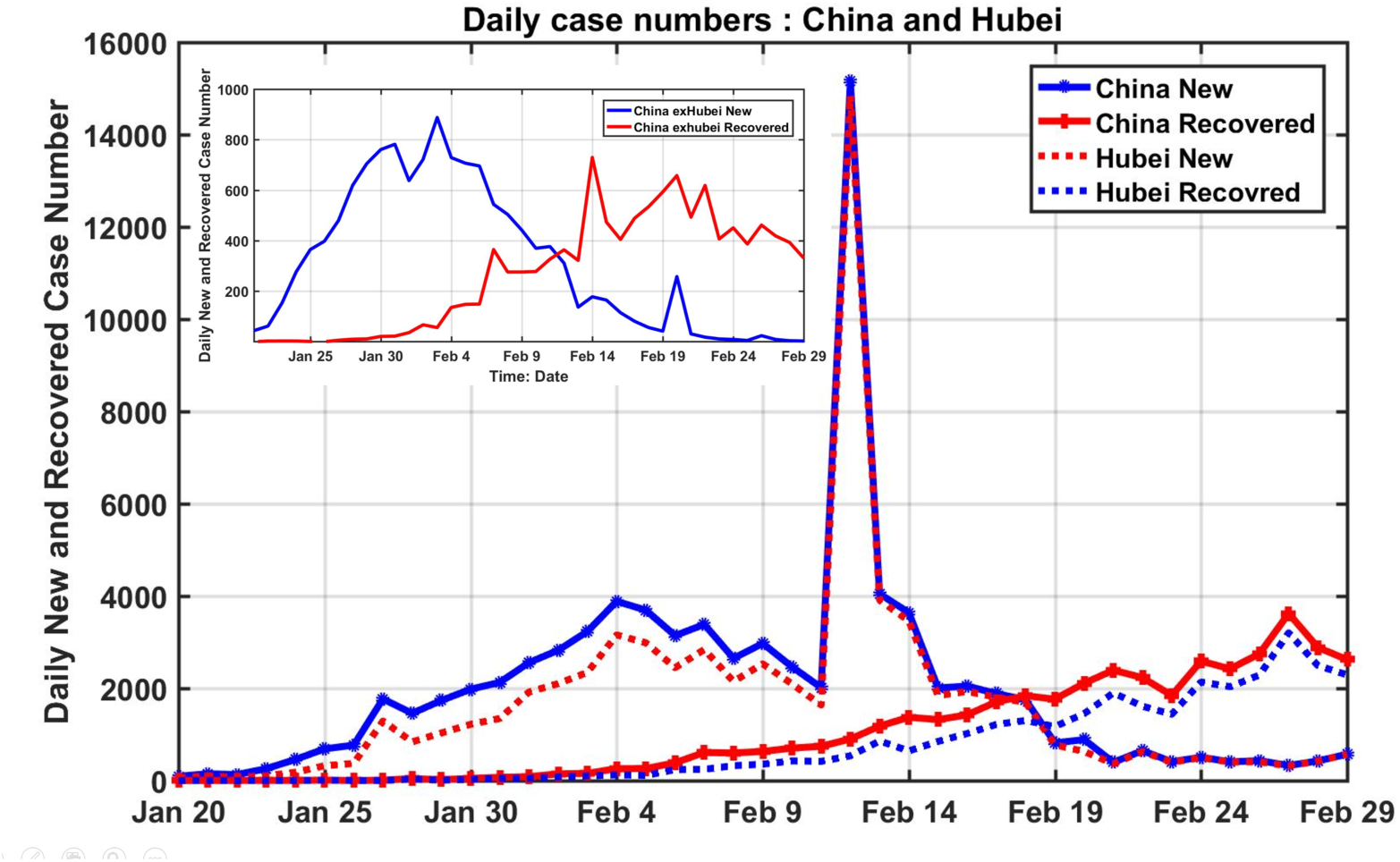
The daily newly infected (in blue] and the daily newly recovered (in red], as a function of time for China as a whole (in solid lines) and Hubei (in dotted red and blue lines). The turning point is determined by when the red and blue curves cross.

## 5. Inferred epidemiological characteristics

Table 1 summarizes predictions for USA, some European countries, South Korea and China. In some countries, one or both of the critical points have just occurred. Even so, it is still difficult to know if one is at the peak without using our prediction procedure. The *TIC ∞* is the Total Infected Cases for the whole epidemic, calculated as twice the value of *TIC* at *t_N_*, and *AIC*_peak_ is the maximum of *AIC* at *t_p_*. Appendix discusses how these quantities are computed/predicted.

## 6. Discussions and Conclusion

There are actually two modeling approaches: mechanistic and statistical models. Mechanistic model is based on precisely defined epidemical parameters. Without *a priori* knowledge of the disease characteristic and existing social contact conditions, however, many of the parameters used are based on assumptions and educated guesses. Nevertheless, they are useful for exploring different scenarios for the policy decision to fight an epidemic prior to the rapid rise of the outbreak. In this paper, we introduced a statistical model that could also serve an important role complementary to the mechanistic models, to be used after the outbreak started. It can provide detailed tracking and prediction of the course of the epidemic. For the statistical model to work well, good quality data are indispensable. Our study indicated that with quality data, tracking and prediction of critical events such as the peak and the turning point could be accurate within days.

Our model is supported by underlying theoretic foundation and validated by the existing data from the region where the pandemic is waning. Many parameters characterizing an epidemic can be determined from local-in-time data. Because it is based on general epidemiological principles we suggest that our approach could be applied not just to the current Covid-19 epidemic, but also generally to future epidemics.

Importantly, we made explicit the concept of “suppressed equilibrium” as an end state of an epidemic in additional to the traditional “herd immunity” state. Based on the traditional mechanistic model, an epidemic ends after a high percentage of the population is infected and recovered hence acquiring immunity. This is the so-called “herd immunity” idea. For COVID-19, which we found to be very contagious, more so than previously thought, the “herd immunity” end would require almost all of the population be infected and therefore would bring unthinkable toll in the number of people sick and dead. A second way for an epidemic to end is with strict contact-reduction measures, so that although a large pool of susceptible population still exists, the portion that an infected person comes in contact with is reduced by the measures adopted, again leading to the Effective Reproductive Number less than 1. Unlike the first end state mentioned above, this “suppressed” state is “parametrically unstable” in the sense that if the social distancing measures are relaxed before the epidemic ends or new infection is imported after the first wave ends, the epidemic will rebound, as a large portion of the population is still susceptible. For this second state to be a stable equilibrium, the social-distancing measures, and quarantine of cross-border visitors need to be maintained until it is clear that the disease has died off. It is this second state that most countries are now aiming for.

The second state will have a much earlier date for the new cases to peak and the epidemic to end. For the US the peak of the newly infected cases is April 7-11. The epidemic is estimated here to end in the first week of June, assuming the current social distancing/stay-at-home measures are maintained till then, and after that date import of infected from abroad is prevented by strict quarantine of visitors. These assumptions now do not look like they would hold as states begin to open their business prematurely. For the US, we predict that the total number of infected cases is 1.5 to 2.4 million, dependent on the assumption. These are symptomatic cases that require hospitalization.

Since it is the goal of most countries to eventually approach the “suppressed equilibrium”, it is important to note that the slowing growth of the incidence (daily newly infected) that is observed is not a function of biology, but is a result of contact-reduction, which is social science. The mechanism of the exhaustion of susceptibles is not relevant anymore as the number of infected is such a very small percentage of the susceptible population. Therefore it is not necessary to use a model, such as SIR or SEIR, to keep track of the change in susceptibles after the start of the outbreak. If these mechanistic models are to be used to track the progress of an outbreak, the infection rate *a* in the models needs to be adjusted constantly, but it is not clear how to adjust it. In any case, the solution is responding to input of changing model parameters, and not to the natural biological evolution built into the model structure, such as recovery and immunity upon recovery.

This is a new way to look at the field of epidemiology. The observed slowing of the growth of incidence and the cresting of the epidemic curve are dominantly the result of a reduction of “infectivity”, as first proposed by John Brownlee one hundred years ago, except that the change is not due to biology— from the “loss of the infecting power on the part of the organism” as he thought, at least not in the case of COVID-19, but as the consequence of reduced contacts among people in lowering the Reproduction Number, which is defined as the number of other people one infected person would infect. This measure is a product of the average number a typical infected person comes in contact with and the probability that a contacted person becomes infected. While the study of the latter is biology and medicine, that for the former is social science and public health.

## Appendices: THEORY

### Definition

Let *I*(*t*) be the number of active infected at time *t*. Its change is given by;

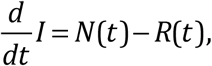

where *N*(*t*) is the number of newly infected, and *R*(*t*) that of the newly recovered or removed (dead). The term: Acting Infected Case (*AIC*) number is used to denote the confirmed *I*(*t*) when we deal with data.

Let *t_p_*, the turning point defined as the peak of the active infected number. At this point maximum medical resource is needed. This maximum occurs when

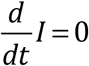 mptymg *N*(*t_p_*) = *R*(*t_p_*).

There is no need to first find *I*(*t*) to locate this peak. Figure 8 shows how this is determined locally. This local-in-time metric avoids the accumulation of poor early data. After the turning point, the newly recovered starts to exceed the newly infected. The demand for medical resources, such as hospital beds, isolation wards and respirators, starts to decrease.

We consider a solitary outbreak. Let *t =* 0 be when the first infection began. For Wuhan, China, this date is near the end of 2019, perhaps even earlier. Let *t_B_* be the beginning of the better quality data. This time is beyond the initial incubation period of the disease and it can be assumed that at that time there is already a population of infected, some of them asymptomatic but nevertheless infectious.

Let *X*(*t,s*) be the number of infected cases at time *t*, withs being the “age” distribution, i.e. number of days sick.

The total number of infected is given by:

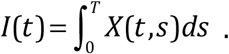

After being sick for *T* days, a patient either recovers or is removed (dead). *T* is called the recovery period (or removal period). It is also called the infectious period if the patient is infectious during this period. Of course its value varies by patient and by the efficacy of treatment for each hospital. For the removed it also depends on the age of the patient and whether there are underlying medical conditions. Only a mean recovery period is obtainable from data, and so this is in reality a statistical quantity. We will discuss later how this statistical quantity can be obtained from data.

### Conservation law

(see ref(22)):

After first infected and until removed or cured, we have:

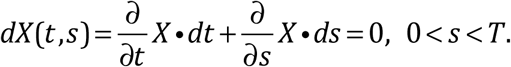

*So*, since *ds* / *dt =* 1,

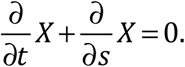

This equation is to be solved using the method of characteristics as *X*(*t*, *s*) = constant along characteristics defined by *ds/dt =* 1.

Boundary condition: *X* (*t*,0), specifies the "birth" process, i.e. how the disease spawns newly infected (with "age" *s =* 0).

Initial condition: X(0, *s*) = 0 for *s >* 0, specifies the initial age distribution at *t =* 0

There are two types of characteristics:

(*i*) *s > t*, (*ii*) *s* < *t*.

The first type of characteristics intersects the *t =* 0 axis, and since the initial condition is zero, we have the solution:

*X*(*t, s*) = 0 for *s* > *t*.

That is, there is no infected population who is sick for more days than the lapsed time since the first infection occurred.

For the second type of characteristics, *t > s* the solution is

*X* (*t, s*) = *f* (*s* −*t*)

with the form of *f* to be determined by the boundary condition. Even without determining the form of *f* we have the following general results:

For *t > T*, and therefore *t > s:*

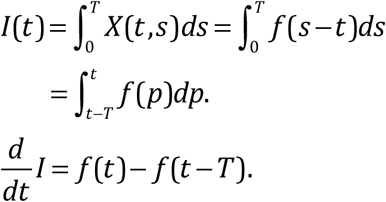

Since the rate of increase of *I*(*t*) is by definition equal to the newly infected number, *N*(*t*), minus the newly recovered (or removed) number, *R*(*t*), we have:

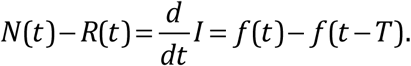

For a fatal disease with low fatality rate, where almost all infected cases eventually recover after a hospital stay of *T* days, we can identify *f* (*t*) with *N*(*t*), and *f* (*t* − *T*) with *R*(*t*).

If the disease has a non-negligible fatality rate, we include the dead in *R*(*t*).

### Main Result

The daily newly recovered/removed number *R*(*t*), is related to the daily newly infected number *N*(*t*) as, for *t > T*:

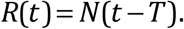

The second type of characteristics intersects the boundary *s =* 0. The boundary condition itself needs to be solved as a function of *t* to describe how new infection (at *s =* 0) occurs. This can be done using a birth model, such as Eq. (1.56) in (23).

*Boundary condition:* Following Murray’s Eq. (1.56), the “birth” is assumed to be proportional to “parents” of suitable “age”, with an age-dependent birth rate, *a*(*s*).

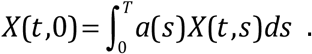

This equation needs to be solved numerically, except in the case of constant *a*. The solution for *X* (*t*, 0) = *g*(*t*) is generally an increasing function of time.

Since *X*(*t,s*) = *g*(*t*−*s*), at some time *t_B_* > *T*: *X*(*t_B,_s*) = *X*(*t_B_*,*s*)= *g*(*t_B_*−*s*)*f*_0_(*s*). That is, an “age” distribution can be converted into a time profile through the method of characteristics.

The solution of the integral equation is not presented here. For our purpose here it suffices to assume that the solution of this model yields a distribution with age that has a full spectrum 0 < *s < T* of infectives at a time *t_B_*, long after a full incubation period has passed.

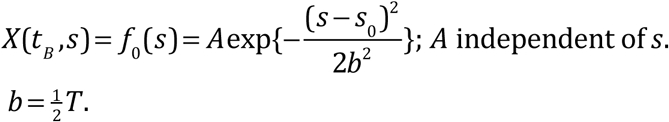

This distribution is justified as follows:

To the right of the peak in “age” distribution, it is easy to understand that the numbers for those who are “older” should be less because they were infected earlier during an earlier stage of the epidemic. As the epidemic grows exponentially, there are more and more “younger” infectives. To the left of the peak in “age’ distribution many are at various stages of incubation. Those who are newly infected may be large in number, but they are less infectious and contribute less to the growth of the subsequent infection, and since they have not developed symptoms, they are less likely to be hospitalized, tested and contribute to the “case” record. For Covid-19, the peak infectiousness occurs in a period just before and after the onset of symptoms, around 5-6 days since first infected (23). The above discussion gives an epidemiological justification for a Gaussian-like distribution of the “age” distribution of the “new cases”, more appropriately called “new hospitalization per day”.

It is important to point out that the assumption of less “cases” to the left of the peak is the only “filter” that serves to limit our theoretical discussion to the data of symptomatic cases. If all infected were included, *f*_0_(*s*) should have a maximum at *s =* 0.

Therefore the solution is, for *t > t_B_ >* 0:

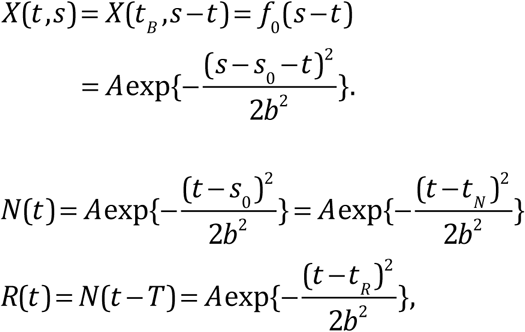

where *t_N_* is the peak of *N*(*t*), and *t_R_ = t_N_ + T* is the peak of *R*(*t*). Both distributions are Gaussians.

For *t_B_ < t < T*,

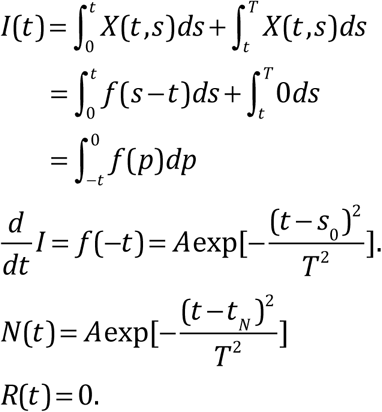

Again, *N*(*t*) is Gaussian, but there is no recovered or removed during this early stage.

### Main Result

The natural logarithm of the ratio of *N* and *R* is a linear function of time for *t > T*:

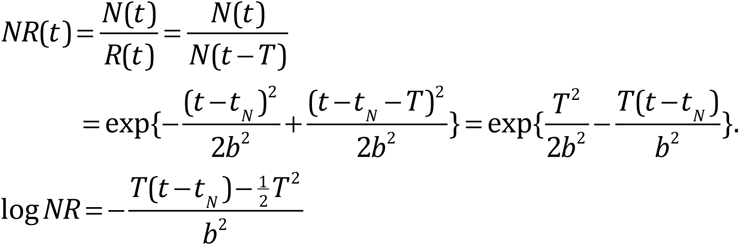

a linear function of *t*.

### Heterogeneous Data

The above results are obtained for the case of a single introduction into a region of infected at *t=*0 and we solve for the subsequent development of the epidemic from that single source. Consider now a large region consisting of a number of small regions, and the “seeding” of the infected occurs at different times for different regions. The large region could be China, and the first infection could be Wuhan, Hubei and then the regions outside Hubei. Then we may have for the China as a whole data for the newly infected a sum of several Gaussians staggered in time. As long as the Gaussians are not separated so much that there are different peaks in the combined data, the combined data can still be considered as Gaussian, as is the case in the real data. However, the standard deviation *σ* of the combined Gaussian is inevitably larger and is no longer given by *b:*

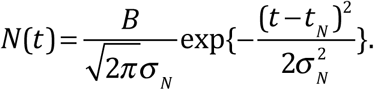

We still have *R*(*T*)=*N*(*t*−*T*) since this result holds for each sub-region. The result that log *NR* is a linear function of time still holds:

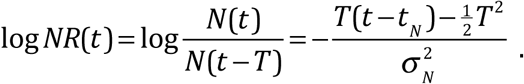

The slope of the straight line is 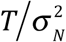

Since the hospital stay can act as a smoothing filter on *N*(*t*) to yield *R*(*t*), the standard deviation for *R*(*t*) could be slightly wider than that for *N*(*t*). So we could have two different Gaussians (but their integral over all time should be the same):

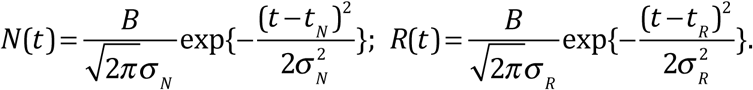

Taking this into account, we have, denoting *T* = *t_R_*−*t_N_*:

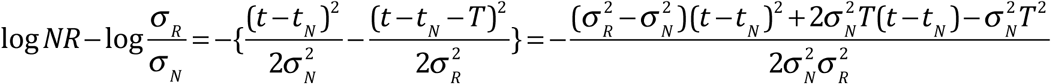

As the values of *σ_N_* and *σ_R_* are very close based on the empirical data, the quadratic term is always small comparing to the other terms for the length of time we are considering here. Hence,

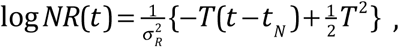

a linear function of time. Its slope is 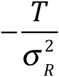.

A caveat: For widely separated countries, such as China and US, the two Gaussian peaks are separated in time. These two regions should be treated separately so that the data for each region has only a single peak. Similarly for Europe and China. See Appendix. There is also a problem with the aggregate data for the US consisting of different states with outbreaks separated in time.

### Result

The natural logarithm of the ratio of two Gaussians of slightly different standard deviation is approximately a straight line, providing the data used is homogeneous temporally and spatially.

Sometimes, heterogeneity in one variable could be relaxed in an overall mean. The case in China is an example. The case could be treated as whole China even though China included Hubei and Wuhan and the very different region of China outside Hubei, for the outbreak occurred at the same time. But the cases in China and European countries and US could not be treated as a whole, for they are separated both spatially and temporally. Consequently, it would be impossible to talk about a single distribution for the global prediction.

### Time-varying *σ* and *t_N_*

The above derivations continue to hold even for *σ* being a function of time. This becomes relevant when, for example, after the epidemic passes its peak the policy makers decide to relax the restrictions on social distancing. Such actions alter the course of the epidemic and create a longer tail of new cases. *N*(*t*) is no longer Gaussian because it does not have fore-aft symmetry. Since such government actions are not included in the initial prediction, the prediction scheme needs to adapt using real-time data after the peak. Our method allows for it.

### The peak infected cases

Writing 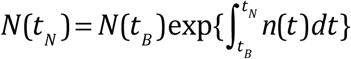, and noting 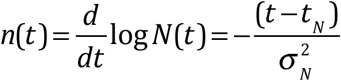, the exponent is 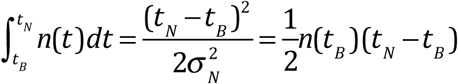. Hence the peak infected case number can be predicted, using the predicted value for *t_N_* starting from a conveniently chosen time *t_B_*, such as the latest time with data available, as:

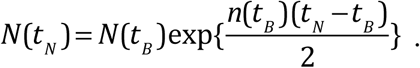

### Accumulated quantities

To calculate *I*(*t*) using reported data, only confirmed cases are used (We call it AIC). It is given by the accumulated newly confirmed cases minus the accumulated confirmed recovered. Since the accumulation of early poor data can introduce errors a more local-in-time formula is given as:

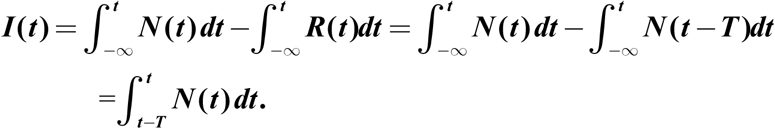

That is, to find *I* at time *t*, one only needs to add up the daily newly infected case numbers for a period of *T* preceding *t*. This is an almost local-in-time property even for this accumulated quantity. For validation, we estimate the peak of the *I* case number on 18 February by computing the sum of daily newly infected case numbers for 15 days, from February 4 to February 18, which yields a peak value for the total infected cases on 18 February of 54,747. This is within 10% of the reported number of 57, 805, even after taking into account the deaths (by subtracting the accumulated deaths of 2,004 from our estimate).

The Total Infected Cases (TIC) is one of the most reported statistics:

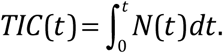

The total infected cases for the epidemic for a region after it is over is given by approximately:

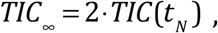

assuming that *N*(*t*) is approximately symmetric about its peak at *t_N_*. In reality *N*(*t*) may not be symmetric and likely has a long tail. However, since the number of cases along the tail is small, the above approximation for the total is still good. Since TIC is officially available at any time *t* we will use that reported number, projected forward to the peak *t_N_*, and then doubling it. Caveat: the official number may include accumulation of early bad data. Hopefully it is a small percentage of the shear size of the TIC.

The peak *AIC* number can be predicted as

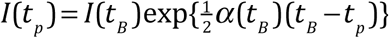, where *t_B_* is the last available data before the turning point. It is assumed that *α*(*t*) lies on a straight line between *t_B_* and *t*_p_. This is the maximum load that health services need to plan for, and adopt “flatten the curve” policies to keep this number under the maximum resources available, such as hospital beds and isolation wards.

### Inset

For China outside Hubei.

## The problem with data quality

The performance of all data-driven models depends on the quality of data. In general, there are problems with data from all countries. Underreporting is universal to all countries at least during the initial period of the epidemic. The causes might be different. In China, for example, the data collection was erratic in the early days for lack of awareness of the novel virus. A more common cause was the shortage of test kits to identify the cases. The situation is made more complicate by the existence of asymptomatic cases. However, whether to include those cases or not depends on the definition. In this study, by definition, the cases are limited to confirmed ones with symptoms, for only those cases would tax the medical resources. In fact, the best any country can do is to record the confirmed new case accurately. The cases for death seem to be simple, for the number of deaths has attracted great attention by the government and public alike. Many models are designed to follow only death cases. However, even here, the numbers are not problem-free. For lack of testing, large number of unattributed COVID-19 deaths exists in every country. Finally, the most critical problem is for the recovered case numbers. It is important to have this record, for it is essential for the conservation law discussed above. Furthermore, the new to recovery/removal case ratio (NR Ratio) provides a robust prediction for the turning point of the epidemic. Unfortunately, the recovered case numbers had attracted the least attention. As a result, it is not recorded or reported in some countries, such as UK. Under this condition, many of the tracking and prediction can still be made as demonstrated in the main text. However, we strongly urge data for the recovery case to be recorded accurately, for it could not only provide accurate prediction for medical resource preparation, but also as a measure of the medical system efficacy.

Finally, a few words on data homogeneity. All models, statistical or mechanistic, require homogeneous condition. There are actually two types of homogeneity: temporal and spatial. We believe the temporal homogeneity is more critical. For example, China can be treated as a whole entity even with part of China, such Hubei and Wuhan under strict lockdown, and part of China is not. The result still makes sense because of its temporal homogeneity. Temporal inhomogeneity, however, would render the data nonsensical. Let us take the global condition as an example. There is a gap in time between the Chinese cases and the rest of the world (see Figure 9). No model should treat the two events as one. See Figure 10. On the other hand, the European country and the US could be treated as the World exChina, for the outbreak in European countries and the US happened at about the same time, even though they are spatially separated widely.

**Figure 9.**
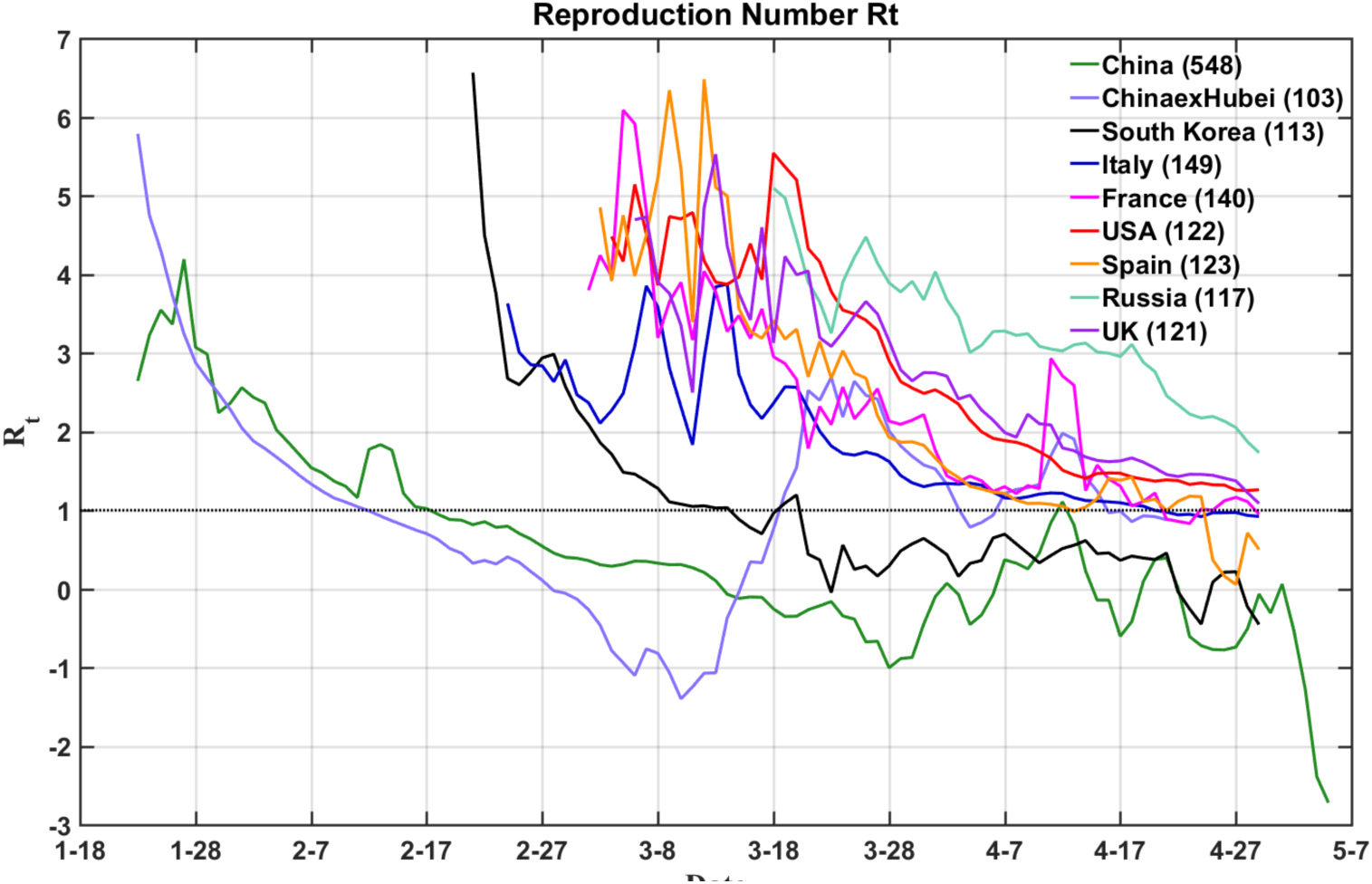
This figure shows the Reproduction Number in real time starting on 20 January. In this presentation, one can see the pandemic is consisted of a serial event with severe data inhomogeneity in both space and time. The outbreaks were almost over in China and South Korea before the earliest cases in Italy and Spain started to rise. There were 40 days difference between China and South Korea on one hand, and the cluster of the pandemic flared up in European countries and the US on the other hand.

**Figure 10.**
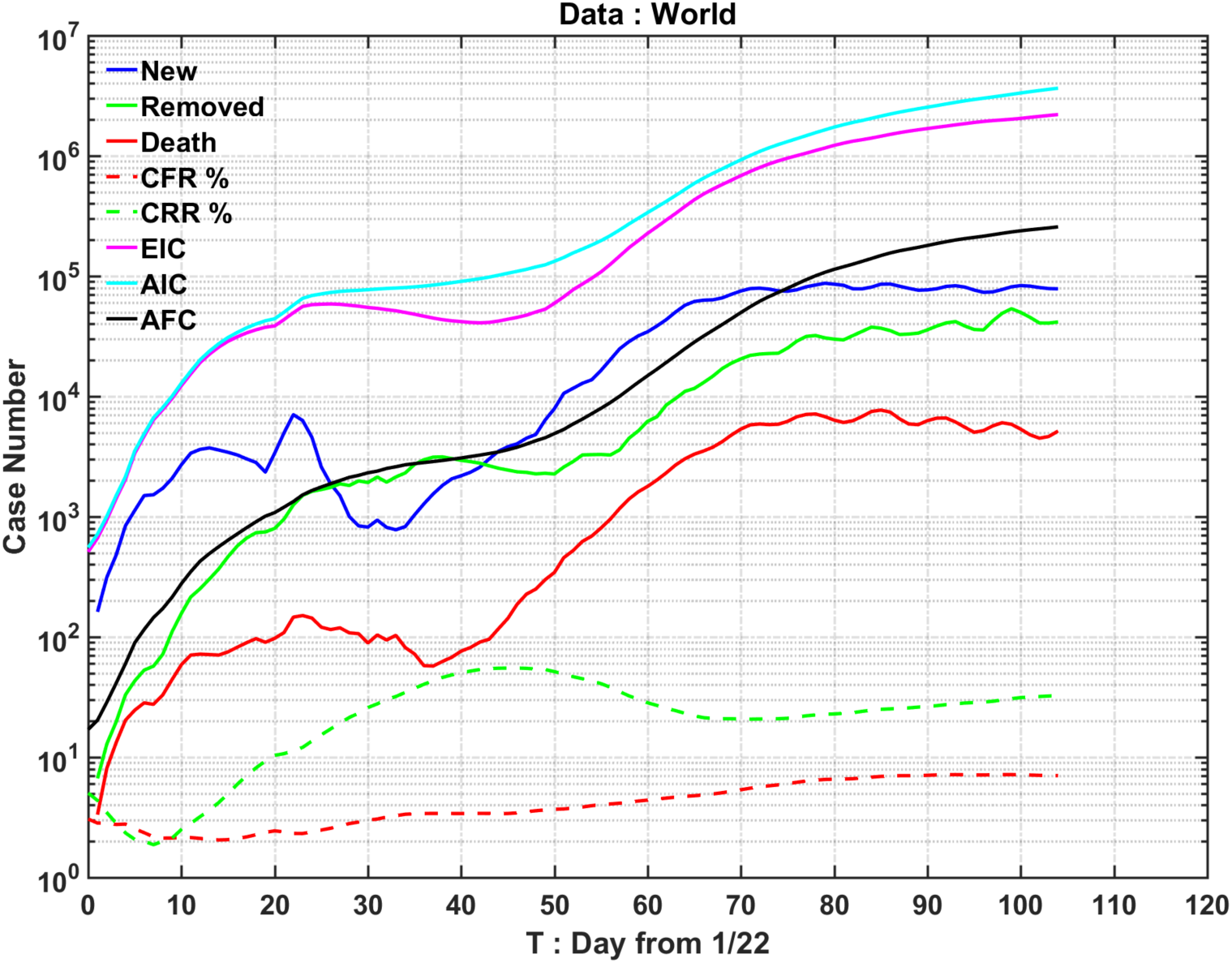
China and the rest of the world should be treat separately.

## Data Availability

All data used in this study are publicly available.

## Acknowledgements

NEH and FQ are supported by the National Natural Science Foundation of China under Grant 41821004. KKT’s research is supported by the Frederic and Julia Wan Endowed Professorship.

## Data Availability

All data in this study are publicly available from World Health Organization (WHO) at https://www.who.int/emergencies/diseases/novel-coronavirus-2019/situation-reports/ and on the Daily Brief site of the China’s National Health Commission at http://en.nhc.gov.cn/ The Korean data is available at https://sa.sogou.com/new-weball/page/sgs/epidemic Coronavirus COVID-19 Global Cases by Johns Hopkins CSSE https://gisanddata.maps.arcgis.com/apps/opsdashboard/index.html#/bda7594740fd40299423467b48e9ecf6

https://github.com/CSSEGISandData/COVID-19/tree/master/csse covid 19 data/csse covid 19 time series

